# Dique Filipeia: A rehabilitation protocol for non-intubated COVID-19 in-hospital patients

**DOI:** 10.1101/2021.07.19.21258787

**Authors:** Murillo Frazão, Kamila Januária de Brito Marinho Paiva, Rossana Maria da Nova Sá, Fábio dos Santos Menezes, Laís Ailenny dos Santos Alves, Anderson Igor Silva de Souza Rocha, Eduardo Eriko Tenório de França, Amilton da Cruz Santos, Maria do Socorro Brasileiro-Santos

**Affiliations:** Municipal Health Department, João Pessoa, PB, Brazil; Laboratory of Physical Training Studies Applied to Health, Federal University of Paraíba (UFPB), João Pessoa, PB, Brazil; Lauro Wanderley University Hospital, UFPB, João Pessoa, PB, Brazil; Physiotherapy Laboratory in Cardiorespiratory Research, Federal University of Paraíba, João Pessoa, PB, Brazil

## Abstract

**Objective:** The aim of this study was to evaluate the effectiveness of the “Dique Filipeia” rehabilitation protocol in patients with COVID-19 admitted to reference hospitals.

**Methods:** This is an experimental study with COVID-19 patients admitted to the hospitals wards being considered eligible. The study outcomes were assessed between patients undergoing the rehabilitation protocol (Dique Filipeia group) and patients who did not receive the protocol (control group). The rehabilitation protocol consisted in classifying patients daily into four levels of severity through peripheral oxygen saturation. Severity was classified by the oxygen flow needed to maintain a saturation greater than or equal to the cut-off point of 93%. A standardized ventilatory support and functional rehabilitation exercises were performed for each severity level patient, followed by an attempt to wean oxygen.

**Results:** A total of 727 patients were analyzed in the study. The Dique Filipeia group presented a lower total (132.7 ± 35.3 vs 307.0 ± 114.3 m^3^/patient; effect size 1.73) and daily (2.9 ± 1.0 vs 6.8 ± 3.1 m^3^/day/patient; effect size 1.46) oxygen expenditure than the control group. The Dique Filipeia patients presented higher hospital discharge (64.9 ± 9.3 vs 35.4 ± 7.5%; effect size 3.46) and lower length of stay (15.8 ± 4.2 vs 29.1 ± 3.4 days; effect size 3.47) than the control group. The Dique Filipeia group patients, who were demanding oxygen therapy, were using 6.2 ± 4.3 L/min of oxygen at day 1. There was a statistically significant reduction from day 2 (p = 0.0001) and oxygen flow was reduced below 1L/min after day 7.

**Conclusions:** The implementation of a standardized rehabilitation protocol reduced oxygen expenditure, increased hospital discharge and reduced the length of hospital stay. Dique Filipeia is a practical, feasible and safe protocol.

## Introduction

The COVID-19 pandemic has promoted a growing amount of cases and deaths in Brazil and worldwide, leading public managers to have great concern and being challenged to reduce the contamination of people, the physical impact of the disease and re-establish and provide the best treatment for patients. In the scenario of long-term hospitalizations and high oxygen expenditure, there was a race against time to open new beds and provide oxygen supplementation to prevent the health system from collapsing. In this sense, it is necessary to propose early rehabilitation protocols which would help in the recovery of patients and therefore lead to greater availability of hospital beds for patients to be treated.

Patients with COVID-19 progress to an inflammatory phase, develop viral pneumonia and may experience dyspnea and hypoxia. Other patients progress to a severe condition which manifests as systemic hyperinflammation syndrome and multiple organ dysfunction(1). It affects several systems and, depending on the degree of evolution, patients with severe end-stage lung disease may require mechanical ventilation.

There is evidence of myocardial damage in the cardiovascular system, with some patients presenting abnormalities similar to myocarditis(2). Furthermore, patients with severe disease have greater neuromuscular affect probably due to myositis and greater inflammatory pattern(3), and activation of inflammatory cytokines causing inflammatory damage to muscle cells(4). In addition to COVID-19, it is known that regardless of any specific pathological process, physical inactivity causes significant muscle fiber atrophy. In this sense, patients with COVID-19 (because they are suitable for a prolonged bed rest) may have atrophy of type I fibers, accompanied by a change from type I and IIa fibers to type IIb fibers. Furthermore, the pathological process in myopathies results in dysfunction and loss of individual muscle fibers located in the motor unit.

As a result of this multisystemic effect which compromises cardiorespiratory fitness and muscle performance in patients with COVID-19(5), (these indicators interfere in the recovery and evolution of the disease), early physical therapy strategies for patients hospitalized in healthcare units are needed to speed up recovery.

With this challenge to organize an effective and feasible rehabilitation protocol, a health team carried out a review of the scientific literature to design a practical and optimized rehabilitation protocol in order to combat the greatest number of physiological changes imposed by the disease(1)(5)(6)(2)(7), which led to a protocol called “Dique Filipeia” being created.

Thus, the aim of this study was to evaluate effectiveness of the Dique Filipeia rehabilitation protocol in patients with COVID-19 admitted to reference hospitals. Our hypothesis is that this protocol will reduce oxygen expenditure, increase hospital discharge and reduce the length of hospital stay.

## Methods

### Study design

This is an experimental study. The Dique Filipeia Protocol was applied in three COVID-19 reference hospitals of the city of João Pessoa, PB, Brazil. The research was coordinated by the General Commission of the Dique Filipeia Protocol by the Laboratory for the Study of Physical Training Applied to Health and by the Laboratory of Physiotherapy in Cardiorespiratory Research, both from the Federal University of Paraíba. This study was approved by the local research ethics committee (Instituto de Educação Superior da Paraíba - IESP, opinion No. 4.777.641, CAAE: 47672821.8.0000.5184) and was registered in the Brazilian Clinical Trial Registration Platform (Number: RBR-107wv6d9).

### Sample

Patients admitted to the hospital wards from March 16^th^ to April 30^th^ 2021 were considered eligible for this study (protocol group). All patients received physiotherapy treatment. Additionally, patients who were admitted to the hospital wards from February 1^st^ to March 15^th^ 2021 were considered in the study as a control group. All patients had COVID-19 diagnosis established by clinical symptoms (fever, fatigue, muscle soreness, cough, dyspnea, etc.) associated with a positive nasal swab test and chest tomography (ground-glass opacity).

### Dique Filipeia protocol

The treatment protocol initially consisted of classifying patients daily into four levels of severity through peripheral oxygen saturation. Severity was classified by the oxygen flow needed to maintain a saturation greater than or equal to the cut-off point of 93%(8).

Level 0: the patient was able to maintain saturation in room air. Level I: the patient was able to maintain saturation at 1 to 2 liters of O_2_ per minute. Level II: the patient was able to maintain saturation at 3 to 4 liters of O_2_ per minute. Level III: the patient needed more than 4 liters of O_2_ per minute to maintain saturation.

After severity classification, a standardized ventilatory support and functional rehabilitation exercises were performed, followed by an attempt to wean oxygen. Severity level classification (and specific treatment) was maintained for 24h, even if oxygen weaning was successful. The oxygen weaning focused on reducing oxygen by at least 1L/min at the end of every physiotherapy session, keeping an oxygen flow to maintain 93% saturation. A detailed description of the protocol according patient severity follows:

#### Level 0

##### Ventilation strategy

Guide the patient to stay in prone position twice a day for at least 30 minutes. No ventilatory support was implemented.

##### Functional rehabilitation strategy

Strength training for lower limbs in closed chain (sitting and standing exercise), 3 sets of 5-10 repetitions (twice a day). Walking for 5 minutes (twice a day). In cases of bed restriction (long-term bedridden, neurological diseases, etc.), neuromuscular electrical stimulation (NMES: pulse width of 400 microseconds, 75Hz frequency, sufficient intensity to promote visible muscle contraction, ON time = 5s and OFF time = 5s) of the quadriceps, hamstrings and anterior tibialis was performed for 15 minutes (once a day) and sitting for 5 minutes (after NMES).

#### Level I

##### Ventilation strategy

CPAP of 7-10cmH_2_O for one hour (twice a day). Oxygen support to maintain peripheral oxygen saturation ≥ 93% during therapy. Guide the patient to stay in prone position twice a day for at least 30 minutes.

##### Functional rehabilitation strategy

Strength training (during CPAP therapy) for lower limbs in closed chain (sitting and standing exercise), 3 sets of 5-10 repetitions (twice a day). Walking for 5 minutes with oxygen support (twice a day). In cases of bed restriction (including not reaching oxygen saturation ≥ 93% during ventilatory strategy), NMES (pulse width of 400 microseconds, 75Hz frequency, sufficient intensity to promote visible muscle contraction, ON time = 5s and OFF time = 5s) of the quadriceps, hamstrings and anterior tibialis was performed for 15 minutes (once a day) and sitting for 5 minutes (after NMES).

#### Level II

##### Ventilation strategy

CPAP of 7-10cmH_2_O for two hours (twice a day) in eupneic patients. Bi-level (IPAP of 14-20cmH_2_O and EPAP of 7-10cmH_2_O, with standard programming of IPAP = 14 cmH_2_O and EPAP = 7cmH_2_O) for two hours (twice a day), in dyspneic patients (dyspnea was established if there was use of accessory muscles and/or nose flapping and/or respiratory rate greater than 24). Oxygen support to maintain peripheral oxygen saturation ≥ 93% during therapy. Guide the patient to stay in prone position twice a day for at least 30 minutes.

##### Functional rehabilitation strategy

Strength training (during CPAP or Bi-level therapy) for lower limbs in closed chain (sitting and standing exercise), 3 sets of 5-10 repetitions (twice a day). Walking for 5 minutes with oxygen support (twice a day). In cases of bed restriction (including not reaching oxygen saturation ≥ 93% during ventilatory strategy), NMES (pulse width of 400 microseconds, 75Hz frequency, sufficient intensity to promote visible muscle contraction, ON time = 5s and OFF time = 5s) of the quadriceps, hamstrings and anterior tibialis was performed for 15 minutes (once a day) and sitting for 5 minutes (after NMES).

#### Level III

##### Ventilation strategy

Bi-level (IPAP of 14-20cmH_2_O and EPAP of 7-10cmH_2_O, with standard programming of IPAP = 14 cmH_2_O and EPAP = 7cmH_2_O) for two hours (three times a day). Oxygen support to maintain peripheral oxygen saturation ≥ 93% during therapy. Guide the patient to stay in prone position three times a day for at least 30 minutes.

##### Functional rehabilitation strategy

Strength training (during Bi-level therapy) for lower limbs in closed chain (sitting and standing exercise), 3 sets of 5-10 repetitions (twice a day). In cases of bed restriction (including not reaching oxygen saturation ≥ 93% during ventilatory strategy), NMES (pulse width of 400 microseconds, 75Hz frequency, sufficient intensity to promote visible muscle contraction, ON time = 5s and OFF time = 5s) of the quadriceps, hamstrings and anterior tibialis was performed for 15 minutes (once a day) and sitting for 5 minutes (after NMES).

### Analyzed variables

For the following variables were collected for analysis: oxygen expenditure per patient (data provided by medicinal gas company) and daily oxygen expenditure per patient (oxygen expenditure per patient divided by number of days of hospital stay), percentage of hospital discharge and length of hospital stay. Oxygen flow need to maintain peripheral oxygen saturation at 93% was only collected in the Dique Filipeia group during a 15-day period.

### Statistical analysis

Data normality was verified using the Shapiro-Wilk test. An unpaired T-test was used to evaluate intergroup differences in age. The categorical variables were analyzed by Fisher’s exact test. The effect size was calculated by the T-test (difference between two dependent means) and post hoc analysis. The input parameters were total sample size, means and standard deviations, and error probability α = 0.05. The effect size points were small = 0.2, medium = 0.5 and large = 0.8 (9). The effect size was used to determine clinically significant differences (large effect size was assumed as clinically significant). An ordinary one-way ANOVA was used to evaluate the intragroup differences. A statistical significance value of p < 0.05 was set for all analyses. GraphPad Prism 7.0 and GPower 3.1.9.7 software programs were used for the statistical analysis. Data are presented as means ± standard deviations and percentages.

## Results

A total of 727 patients were analyzed in the study. Characteristics of each group are presented in Table 1. Age and sex characteristics, main comorbidities and main drug therapy were similar in both groups.

**Table 1.** Age and sex characteristics, main comorbidities and main drug therapy.

| **Variable** | **Control**<br>**(n = 172)** | **Dique Filipeia**<br>**(n = 555)** | **P value** |
| --- | --- | --- | --- |
| **Age, years** | 53.4 ± 6.4 | 52.3 ± 7.2 | 0.0730 |
| **Sex, M/F%** | 57.5/42.5 | 53.3/46.7 | 0.3833 |
| **Comorbidities, n(%)** |  |  |  |
| Obesity | 29 (16.9) | 117 (15.5) | 0.6321 |
| Hypertension | 59 (34.3) | 214 (38.5) | 0.2126 |
| Diabetes | 52 (30.2) | 145 (26.1) | 0.3261 |
| **Drug therapy, n(%)** |  |  |  |
| Antibiotics | 172 (100) | 555 (100) | 1.0 |
| Corticosteroids | 172 (100) | 555 (100) | 1.0 |
| Anticoagulants | 172 (100) | 555 (100) | 1.0 |

M: male and F: female. Mean ± standard deviation.

The Dique Filipeia patients presented lower total (132.7 ± 35.3 vs 307.0 ± 114.3 m^3^/patient) and daily (2.9 ± 1.0 vs 6.8 ± 3.1 m^3^/day/patient) oxygen expenditure than control, with a large effect size (Figure 1).

**Figure 1:**
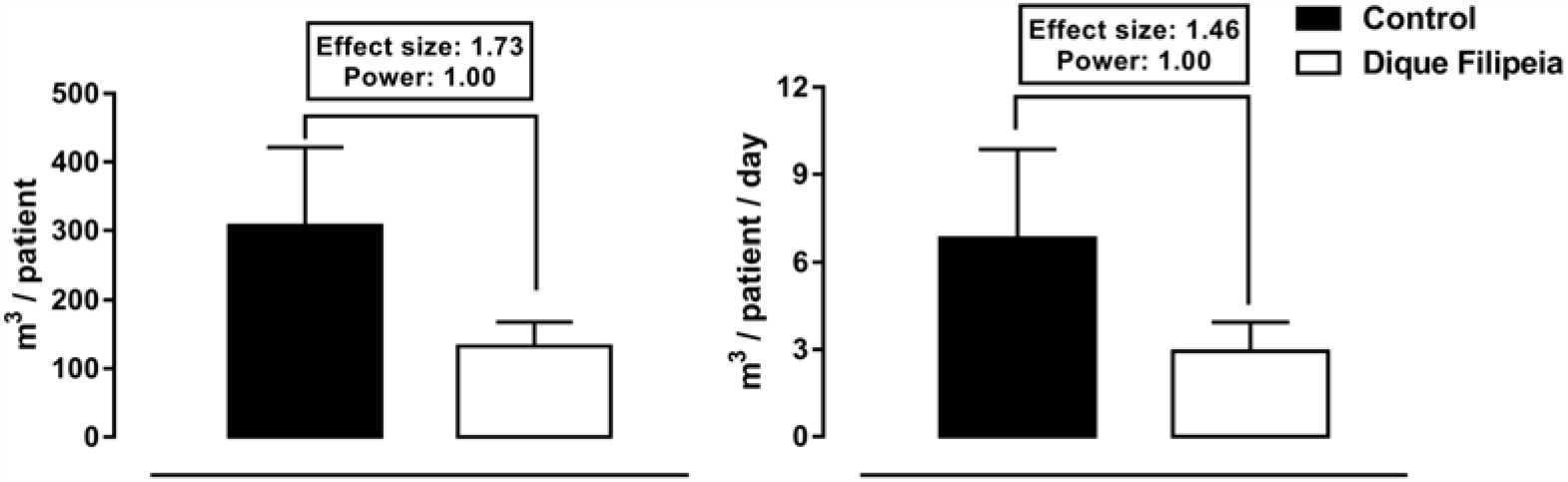
Total (left panel) and daily (right panel) oxygen expenditure. Mean ± standard deviation.

The Dique Filipeia patients presented a higher hospital discharge (64.9 ± 9.3 vs 35.4 ± 7.5%) and a lower length of hospital stay (15.8 ± 4.2 vs 29.1 ± 3.4 days) than the control group, with a large effect size (Figure 2).

**Figure 2:**
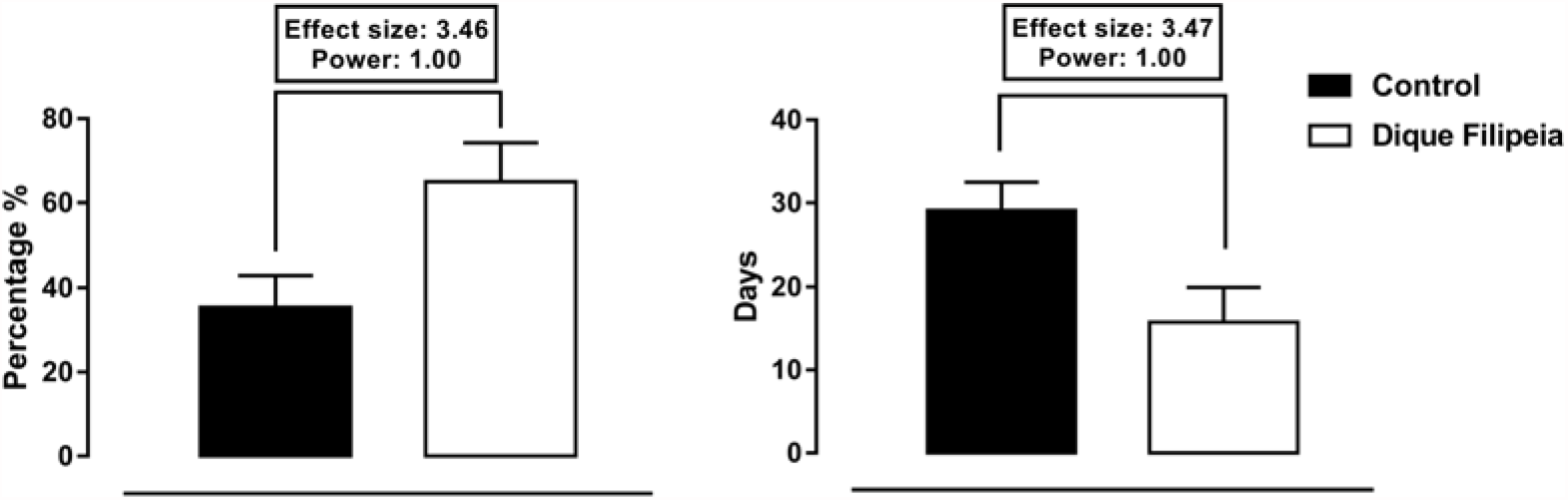
Hospital discharge (left panel) and length of hospital stay (right panel). Mean ± standard deviation.

The Dique Filipeia patients (who were demanding oxygen therapy) were using 6.2 ± 4.3 L/min of oxygen at day 1. There was a statistically significant reduction starting from day 2 (p = 0.0001) and oxygen flow was reduced below 1L/min after day 7. A stratified analysis showed that Level I patients were using 1.6 ± 0.5 L/min of oxygen at day 1, and oxygen flow was reduced below 1L/min after day 7. Level II patients were using 3.4 ± 0.6 L/min of oxygen at day 1. There was a statistically significant reduction starting from day 2 (p = 0.0004), and oxygen flow was reduced below 1L/min after day 4. Level III patients were using 9.2 ± 3.6 L/min of oxygen at day 1. There was a statistically significant reduction starting from day 3 (p < 0.0001), and oxygen flow was reduced below 1L/min after day 12 (Figure 3).

**Figure 3:**
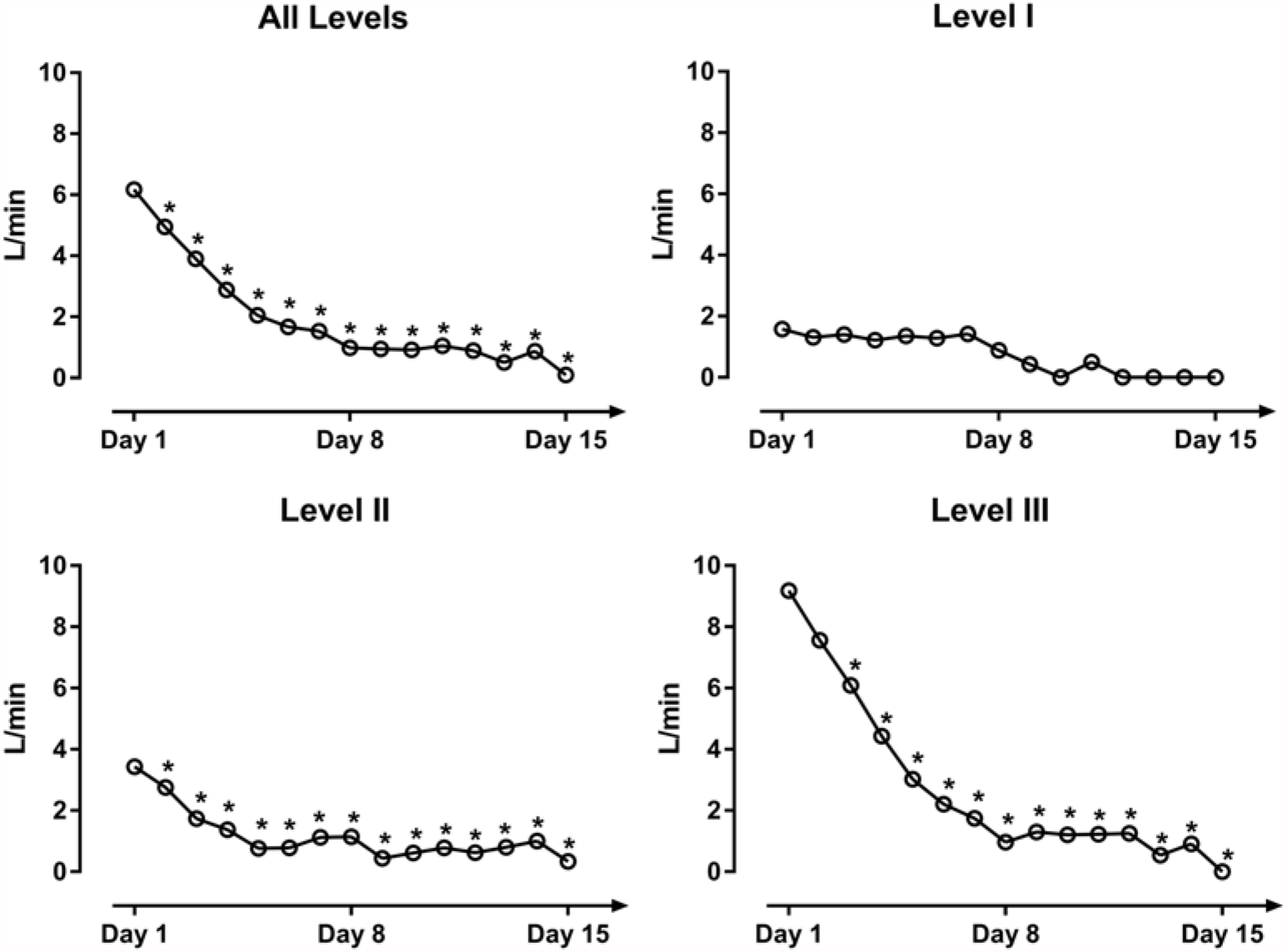
Dique Filipeia patients oxygen weaning. Mean. ^*^ p < 0.01 compared to day 1.

## Discussion

The rationale during the protocol elaboration was that it could help to avoid the collapse of the health system by reducing oxygen expenditure and hospitalization time, providing available beds. The main finding of this study was that the Dique Filipeia protocol: 1) reduced oxygen expenditure; 2) increased hospital discharge; and 3) reduced the length of hospital stay.

Oxygen expenditure reduction was probably reached due to two distinct factors: strict management of oxygen flow to avoid waste and improvement ventilation/perfusion relationship. Oxygen waste is a common practice in Brazil, with patients under a much higher oxygen flow than physiologically needed. There were at least 3 daily oxygen saturation monitoring moments during the protocol application. The oxygen flow was strictly set to maintain 93% oxygen saturation. Moreover, non-invasive ventilation (NIV), prone position and low to moderate intensity exercise were used to improve the ventilation/perfusion ratio.

NIV increases alveolar pressure, recruiting alveolar units and putting them closer to capillary bed flow, improving oxygenation. In a retrospective study with 222 patients, Daniel et al.(10) demonstrated that utilization of NIV as the initial intervention in COVID-19 patients requiring ventilatory support was associated with a significant survival increase. The mortality rate for patients intubated after NIV is not worse than those who undergo intubation as their initial intervention(10). The Dique Filipeia NIV strategy increases ventilatory support parameters and time utilization according to patient severity. Our perception is that these individual adjustments were essential to therapy success.

The prone position is related to a more homogeneous distribution of ventilation(11). It improves oxygenation by reducing lung ventilation/perfusion mismatch and promoting recruitment of non-aerated dorsal lung regions. Vittorio et al.(12) demonstrated that the prone position improves oxygenation in spontaneously breathing non-intubated patients with hypoxemic acute respiratory failure. Unfortunately, these benefits are not maintained for a long time. That was the reason to repeat the maneuver 2-3 times a day.

Pulmonary vascular performance is deeply impaired by COVID-19. A hypercoagulable state associated with COVID-19 results in micro and macrothrombosis of the pulmonary vasculature(13). Ventilation/perfusion mismatch is increased (increased dead space) and hypoxemia is worsened. The Dique Filipeia strategy was implemented to avoid/reduce the risk of coagulation. Exercise has been demonstrated to have considerable effects on hemostasis by modifying coagulation and fibrinolytic markers. Low to moderate intensity exercise is likely to reduce the risk of coagulopathy response(14).

Another Dique Filipeia strategy was to acutely improve pulmonary blood flow to reduce ventilation/perfusion mismatch. It has been shown that exercise is normally associated with a decrease in pulmonary vascular resistance. The exercise-induced decrease in pulmonary vascular resistance is attributable to pulmonary vascular recruitment in setting increased pulmonary arterial pressure and flow, as well as the distensibility of the normal pulmonary resistance vessels. Mild to moderate exercise enables complete pulmonary vascular recruitment(15).

The major limitations to exercise (hypoxemia and dyspnea) were resolved by implementing oxygen and ventilatory support during low to moderate intensity exercise. It is important to emphasize that patients only performed the exercises in a safe condition (oxygen saturation > 93% and absence of dyspnea). If the safe condition was not reached, patients did not perform the exercise protocol. NIV and oxygen support reduces breathing work and respiratory muscles’ demand during exercise, increasing performance. O’Donell et al.(16) demonstrated that improvement is primarily explained by altered symptoms (reducing dyspnea and leg discomfort) and did not directly correlate with alterations in any of the measured cardiopulmonary physiological variables.

Hospital discharge improvement and length of hospital stay reduction was achieved due to the combination of progressive oxygen weaning and the maintenance of the performance of physiological systems. Cardiorespiratory fitness and neuromuscular performance are directly affected by severe COVID-19(5). Exercise training helps keeping the physiological gears running. It prevents cardiac atrophy during the bed rest period(17), improving cardiovascular performance. It also improves strength and neuromuscular efficiency, even at moderate intensity(18), and reduces inflammatory markers(19). In patients who are not able to perform voluntary muscle contraction, neuromuscular electrical stimulation prevents muscle atrophy(20) and acutely mobilizes endothelial progenitor cells (maintaining muscle perfusion)(21).

Exercise may also have a silver bullet for COVID-19 treatment, as irisin concentration substantially increases(22) immediately after an acute bout of exercise. Irisin is a myokine secreted by myocytes during muscle contraction. Oliveira et al.(23) found a positive effect of irisin on the expression of multiple genes related to viral infection by SARS-CoV-2. Irisin diminishes genes associated to viral replication improvement and increases genes associated to viral replication reduction.

Performing the Dique Filipeia protocol was feasible and safe. Temporary desaturation (below 90%) only occurred in 25 (4.5%) patients during first set of sitting and standing exercise. It was quickly reverted after oxygen support adjustment. No major adverse events (irreversible desaturation, irreversible dyspnea, hemodynamic instability, etc.) were registered during protocol execution.

This study has some limitations. We have no detailed data about oxygen therapy guidance, ventilatory support or physiotherapy protocols before the Dique Filipeia period (the physiotherapy treatment was not standardized in these hospitals). We had no data to compare the oxygen flow needed to maintain peripheral oxygen saturation at 93% or the level of patient severity before the Dique Filipeia period.

## Conclusions

The implementation of a standardized rehabilitation protocol reduced oxygen expenditure, increased hospital discharge and reduced the length of hospital stay. The Dique Filipeia protocol is practical, feasible and safe.

## Data Availability

All data presented in this manuscript is available.

